# T helper-1 activation via interleukin-16 is a key phenomenon in the acute phase of severe, first-episode major depressive disorder and suicidal behaviors

**DOI:** 10.1101/2023.04.16.23288643

**Authors:** Abbas F. Almulla, Ali Abbas Abo Algon, Chavit Tunvirachaisakul, Hussein K. Al-Hakeim, Michael Maes

## Abstract

**Background:** Immune-inflammatory pathways in major depressive disorder are confined to the major dysmood disorder (MDMD) phenotype (Maes et al., 2022). No studies have addressed the immune profile of first episode MDMD (FE-MDMD).

**Methods:** This study examines 48 cytokines/chemokines/growth factors, and classical M1, alternative M2, T helper (Th)-1, Th-2, and Th-17 phenotypes, immune-inflammatory response system (IRS), compensatory immunoregulatory system (CIRS), and neuro-immunotoxicity profiles in the acute phase of FE-MDMD (n=71) versus healthy controls (40).

**Results:** FE-MDMD patients show significantly activated M1, M2, Th-1, IRS, CIRS, and neurotoxicity, but not Th-2 or Th-17, profiles compared to controls. FE-MDMD is accompanied by Th-1 polarization, while there are no changes in M1/M2 or IRS/CIRS ratios. The top single indicator of FE-MDMD was by far interleukin (IL)-16, followed at a distance by TRAIL, IL-2R, tumor necrosis factor (TNF)-β. The severity of depression and anxiety was strongly associated with IRS (positively) and Th-2 (inversely) profiles, whereas suicidal behavior was associated with M1 activation. Around 56-60% of the variance in depression, anxiety, and suicidal behavior scores was explained by IL-16, platelet-derived growth factor (PDGF) (both positively), and IL-1 receptor antagonist (inversely). Increased neurotoxicity is mainly driven by IL-16, TNF-α, TRAIL, IL-6 and chemokine (CCL2, CCL11, CXCL1, CXCL10) signaling. Antidepressant-treated patients show an increased IRS/CIRS ratio as compared with drug-naïve FE-MDMD patients.

**Conclusions:** FE-MDMD is accompanied by positive regulation of the IRS mainly driven by Th-1 polarization and T cell activation (via binding of IL-16 to CD4), and TNF, chemokine, and growth factor signaling.

## Introduction

Maes et al. (1990-1995) discovered that major depressive disorder (MDD) is characterized by T cell activation with elevated levels of T helper (Th)-1 products such as serum interleukin-2 (IL-2), soluble IL-2 receptor (sIL-2R), either in serum or culture supernatant of stimulated blood, and stimulated production of interferon (IFN)-γ (Maes and Carvalho 2018). T cell activation in MDD was demonstrated by an increase in the expression of CD4+, CD4+/CD7+CD25+, and CD4+HLA-DR+ cell surface markers using flow cytometry (Maes, Bosmans et al. 1990, Maes, Lambrechts et al. 1992, Maes, Stevens et al. 1993). Moreover, Maes’s laboratories demonstrated that MDD is associated with increased M1 macrophage activation, as evidenced by increased polyclonally stimulated production of IL-1β and IL-6 (Maes, Bosmans et al. 1991, Maes, Scharpé et al. 1993) and increased serum IL-6 and tumor necrosis factor (TNF)-α levels (Maes, Smith et al. 1995, Mikova, Yakimova et al. 2001). Results demonstrating that MDD is associated with elevated serum levels of positive acute phase proteins and complement factors as well as decreased levels of negative acute phase proteins, suggest a mild inflammatory response (Maes 1993).

Numerous reviews and meta-analyses have demonstrated that MDD is accompanied by Th-1 activation and a mild chronic inflammatory process and other aberrations in Th-2 (e.g, increased serum IL-4), Th-17 (increased serum IL-17), and T regulatory (Treg) (increased serum IL-10) phenotypes (Haapakoski, Mathieu et al. 2015, Köhler, Freitas et al. 2017, Maes and Carvalho 2018, Almulla, Thipakorn et al. 2022, Almulla, Thipakorn et al. 2023). Thus, both the immune-inflammatory response system (IRS) (activated M1, Th-1, Th-17) and the compensatory immunoregulatory system (CIRS) (Th-2, Treg) may be activated in MDD (Maes and Carvalho 2018). The CIRS exerts negative immune-regulatory functions in an effort to prevent hyperinflammation.

It should be emphasized that the aforementioned meta-analyses were based on serum levels of immune-inflammatory mediators, whereas our early findings also demonstrated elevated levels of stimulated IL-1β and IFN-γ production in MDD (Maes, Bosmans et al. 1991, Maes, Scharpé et al. 1994). The levels of cytokines in the culture supernatant of polyclonally stimulated whole blood or peripheral blood mononuclear cells (PBMCs) reflect the in vivo situation, and additionally indicate whether the immune system is sensitized to respond to immune injuries (Maes, Nani et al. 2021).

It should be stressed that both the serum levels of selected M1/Th-1 products (e.g., IL-1, sIL-1RA, IL-6, TNF-α, and neopterin) (Celik, Erdem et al. 2010, Maes, Mihaylova et al. 2012, Sowa-Kućma, Styczeń et al. 2018, Maes, Moraes et al. 2019) and the stimulated production of IRS and CIRS profiles (Maes, Rachayon et al. 2022) are affected by the recurrence of illness (ROI), defined as the number of depressive episodes and suicidal attempts (Maes, Rachayon et al. 2022). However, only limited information is available regarding isolated cytokines/chemokines/growth factors in the first episode of MDD (FED). Zou et al. reported aberrant cytokine levels in drug-naïve MDD patients (Zou, Feng et al. 2018) and Tang et al. found that TNF-α and IL-10 were elevated (Tang, Liu et al. 2021). IL-6, TNF-α, CRP, IL-1β, sIL-1RA and IL-2 were significantly elevated in drug-naive FED (Çakici, Sutterland et al. 2020, Yang, Liu et al. 2021), whereas IL-8 was significantly decreased (Çakici, Sutterland et al. 2020).

Using machine learning techniques, a new discovery was made that there are in fact two types of MDD, namely major dysmood disorder (MDMD) and simple DMD (SDMD) (Maes, Moraes et al. 2021, Maes, Rachayon et al. 2022). The first phenotype is characterised by increased severity of depression, anxiety, fatigue, suicidal behaviors (SB), and physio-somatic symptoms, lowered quality of life, more cognitive dysfunctions and disabilities; and disorders in (auto)immune, oxidative stress, antioxidant defenses, and gut microbiota that are not detected in patients with SDMD (Maes, Moraes et al. 2021, Simeonova, Stoyanov et al. 2021, Maes 2022, Maes, Rachayon et al. 2022, Maes, Vasupanrajit et al. 2023). Importantly, the stimulated production of M1, Th-1, Th-2, Th-17, Treg, IRS, CIRS, and growth factors, was significantly higher in MDMD than in controls, while no such disorders could be established in SDMD (Maes, Rachayon et al. 2022). Importantly, some M1 (IL-1β, IL-6, IL-15, TNF-α, CCL2, CCL5, CXCL10), Th-1 (IL-2, IFN-γ, IL-16), and Th-17 (IL-6 and IL-17) and even Th-4 (e.g. IL-4) products have neurotoxic effects by damaging neuronal functions that regulate affection, leading to neuro-affective toxicity and, thus, depression (Maes, Rachayon et al. 2022).

Nevertheless, there are no data on the serum IRS and CIRS immune profiles and serum cytokines/chemokines/growth factors of first-episode MDMD (FE-MDMD), nor do we know if FE-MDMD is characterised by an increase in M1/M2 or Th-1/Th-2 polarisation, and an increase in the IRS/CIRS ratio. Such information is crucial because the specific disorders identified in FE-MDMD are the primary drug targets for treating the disorder and preventing future episodes. Consequently, the purpose of this investigation is to a) compare M1, Th-1, Th-2, Th-17, Treg, IRS, CIRS, neurotoxicity profiles, M1/M2 and Th-1/Th-2 polarization, and single cytokines/chemokines/growth factors in FE-MDMD versus normal controls; and b) delineate the immune and cytokine/chemokine/growth factor profiles of the severity of FE-MDMD and suicidal behaviors.

## Materials and methods

### Participants

For this case-control study, we recruited 71 patients with FED from the psychiatric unit at Alhakeim Hospital in Najaf, Iraq, from October 2021 to March 2022. All of the patients were diagnosed by a senior psychiatrist as having MDD using the Diagnostic and Statistical Manual of Mental Disorders (DSM-5) (American Psychiatric Association 2013). All FED patients were in the acute stage of the illness, and none showed signs of complete or partial remission. The MDD patients fulfilled the criteria to be categorized as MDMD (Maes, Vasupanrajit et al. 2023). This diagnosis can be made using machine learning models that we constructed based on the Hamilton Depression Rating Scale (HAMD) (Hamilton 1960) and Hamilton Anxiety Rating Scale (HAMA) (Hamilton 1959) rating scores, and SB and ROI scores or when the HAMD score is ≥ 22 and HAMA score ≥ 22. All patients included showed HAMD and HAMA values that fulfilled the criteria of first episode MDMD (FE-MDMD). The mean duration of illness for patients with FE-MDMD is ±2.5 months. Moreover, the same senior psychiatrist also recruited 40 healthy control individuals from the same catchment region as patients’ friends, and medical staff or friends of medical staff.

We omitted participants who met the criteria for the following axis-1 DSM-5 disorders: dysthymia, schizophrenia, schizo-affective disorder, bipolar disorder, autism or autism spectrum disorder, substance use disorder (except nicotine dependence), post-traumatic stress disorder, psycho-organic disorders, generalized anxiety disorder, and obsessive-compulsive disorder. Furthermore, controls were not enrolled if they had ever been diagnosed with major depression or dysthymia or had a family history of depression, mania, psychosis, substance use disorder, or suicide. The study did not include people with chronic liver or kidney illness, and pregnant and lactating women. Stroke, multiple sclerosis, Parkinson’s disease, and Alzheimer’s disease were all disqualifying conditions, as were (auto)immune diseases like psoriasis, rheumatoid arthritis, inflammatory bowel disease, cancer, type 1 diabetes, scleroderma, acute COVID-19 infection, a COVID-19 infection the 6 months prior to enrollment, any severe or critical COVID-19 disease, and Long COVID. Moreover, individuals receiving treatment with immunosuppressive or immunomodulatory medications or therapeutic doses of antioxidants or omega-3 supplements were deemed ineligible for enrollment in the study.

Before participating in the research, all subjects, or their respective parents/legal guardians, provided written informed consent. The study was approved by the ethics committee (IRB) of the College of Medical Technology at the Islamic University of Najaf, Iraq, under (Document No. 18/2021). The research was conducted in compliance with both Iraqi and international ethical and privacy regulations, including but not limited to the World Medical Association’s Declaration of Helsinki, the Belmont Report, the CIOMS Guideline, and the International Conference on Harmonization of Good Clinical Practice. Moreover, our institutional review board strictly adheres to the International Guideline for Human Research Safety (ICH-GCP).

### Clinical assessments

A senior psychiatrist conducting a semi-psychiatric interview determined the sociodemographic, clinical and psychological data using a semi-structured interview. The total scores on the HAMD and HAMA were employed to delineate the severity of depression and anxiety, respectively. Suicidal behaviors were assessed using two items of the Columbia Suicide Severity Rating Scale (C-SSRS), namely a) number of suicidal attempts in the year prior to inclusion in the study, and weekly frequency of suicidal ideation the last three months (Posner, Brown et al. 2011). We computed a z unit-weighted composite score as the sum of the z values of the HAMD suicide item + number of suicidal attempts + frequency of suicidal ideation, labeled SB (suicidal behaviors). Body mass index (BMI) was determined by dividing each participant’s kilogram body weight by their height (squared meters).

### Biochemical assays

We obtained 5 ml of fasting venous blood in the early morning (8:00-11:00 a.m.) from all subjects enrolled in the current study using a disposable syringe and serum tubes. After centrifuging the blood at 35,000 rpm, serum was separated and kept in Eppendorf tubes at -80 °C in small aliquots until they were thawed for biomarker assays. The fluorescence intensity (FI), including the blank subtracted IF, and concentrations of 48 cytokines/chemokines/growth factors were measured using Bio-Plex Multiplex Immunoassay kits (Bio-Rad Laboratories Inc., Hercules, USA). **Electronic Supplementary File (ESF), Table 1** lists all analytes determined in our study and shows the gene ID and aliases as well. Briefly, the procedure consisted of the following steps: a) diluting the serums using sample diluent (H.B.) (1:4), b) the diluted samples were added to a 96-well plate containing 50µl of microparticle cocktail (including cytokine/chemokines/growth factors) per well and incubated at room temperature with shaking at 850 rpm for 1 hour, c) after three rinses, the plate was added to, and incubated with, 50µl of diluted Streptavidin-PE in each well (shaking at 850 rpm) at room temperature for 10 minutes, d) subsequently, 125 µl of assay buffer was added to each well, and the samples were shaken at room temperature (850 rpm) for 30 seconds. The subsequent analysis employed the Bio-Plex® 200 System (Bio-Rad Laboratories, Inc.) for the assessment of the cytokines/chemokines/growth factors. For all analytes, the intra-assay CV values were below 11.0%.

**Table 1:** Sociodemographic and clinical features of patients with first-episode major dysmood disorder (FE-MDMD) and healthy controls (HC).

| **Variables** | **HC**<br>**(n=40)** | **FE-MDMD**<br>**(n=71)** | **F/X<sup>2</sup>** | **df** | **p-value** |
| --- | --- | --- | --- | --- | --- |
| Age (Years) | 32.1 (8.2) | 31.2 (8.6) | 0.28 | 1/109 | 0.600 |
| Gender (Male/Female) | 23/17 | 41/30 | 0.00 | 1 | 0.980 |
| Body Mass Index (m<sup>2</sup>/Kg) | 25.94 (4.18) | 24.30 (3.49) | 4.89 | 1/109 | 0.029 |
| Marital state (Yes/No) | 21/19 | 26/45 | 2.64 | 1 | 0.104 |
| Smoking (Yes/No) | 19/21 | 17/54 | 6.48 | 1 | 0.011 |
| Literate (Yes/No) | 28/12 | 52/19 | 0.13 | 1 | 0.715 |
| Previous COVID infection (Yes/No) | 17/23 | 27/44 | 0.21 | 1 | 0.644 |
| Duration of illness (Months) | - | 2.52 (2.84) | - | - | - |
| HAMD | 3.2 (2.5) | 28.3 (4.3) | MWUT | - | <0.001 |
| HAMA | 2.7 (1.8) | 27.8 (3.7) | MWUT | - | <0.001 |
| Suicidal behaviors | -1.168 (0.115) | 0.658 (0.586) | MWUT | - | <0.001 |

HAMD and HAMA: total Hamilton Depression and Anxiety Rating Scale scores, respectively.

Based on the standard concentrations provided by the manufacturer, we obtained the concentrations and evaluated the percentage of the concentrations greater than the lowest measurable concentration (out of range, OOR). ESF, Table 1 lists the number of analytes that were measurable (values higher than OOR). In the statistical analysis, FI values are employed rather than absolute concentration since FI values are more suitable than absolute concentrations, particularly when using numerous plates (Rachayon, Jirakran et al. 2022). Our statistical analysis excluded cytokines/chemokines/growth factors with concentrations that were out of range in more than 80% of the instances. As such, IFN-α2, IL-3, IL-7 and IL-12p40 were excluded. Analytes with quantifiable levels ranging from 20 to 40% were treated in regression analysis as prevalences (dummy variables). As such, IL-17 was introduced as a dummy variable. However, we also computed various immune profiles (see Introduction) that make use of all analytes, with the exception of those with < 20% detectable concentrations (Maes, Rachayon et al. 2022, Thisayakorn, Thipakorn et al. 2022)].

The primary outcome data in the present study are the immune profiles built using z-unit-weighted composite scores of different cytokine/chemokine/growth factor. ESF, Table 2 shows the variables that were used to construct M1, M2, M1/M2 (z M1 – zM2), Th-1, Th-2, Th-1/Th-2 (z Th-1 – z TH-2), IRS, CIRS, IRS/CIRS (z IRS – z CIRS), and neurotoxicity profiles. If the immune profiles showed significant differences among the groups, we also entered the single cytokines/chemokines/growth factors.

**Table 2:**
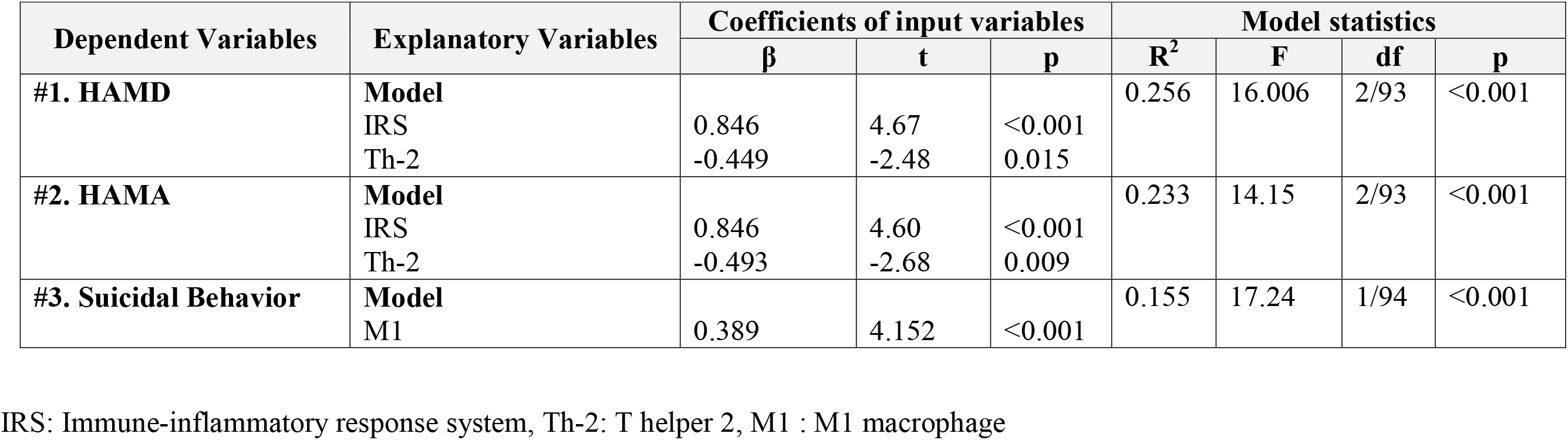
Results of multiple regression analyses with Hamilton Depression Rating Scale (HAMD), Hamilton Anxiety Rating Scale (HAMA) and suicidal behaviors as dependent variables and immune profiles as explanatory variables.

### Statistics

Across study groups, we compared continuous variables using analysis of covariance (ANCOVA) and nominal variables using analysis of contingency tables (χ2-test). We used Pearson’s product moment correlation coefficients to investigate the relationships between scale variables. The impacts of explanatory variables (e.g., cytokine profiles) on dependent variables (e.g., HAMD, HAMA, and SB) were evaluated using manual multiple regression analysis. We also employed an automatic forward stepwise regression approach, with thresholds of p=0.05 to enter and p=0.06 to eliminate, to determine which variables would be included and which would be left out of the final regression model. For all of the explanatory variables in the final regression models, we calculated the standardized coefficients with t-statistics and exact p-values, F statistics (and p values) and total variance (R^2^ or partial Eta squared used as effect size). By utilizing the tolerance (cut-off value: 0.25) and variance inflation factor (cut-off value: > 4), as well as the condition index and variance proportions from the collinearity diagnostics table, we were able to examine the possibility of multicollinearity and collinearity. Heteroskedasticity was confirmed using the White and modified Breusch-Pagan tests. When analyzing the single cytokine/chemokine/growth factor data, we first analyzed the data using forward stepwise automatic linear modeling analyses with the best subsets with overfit prevention criterion or with the overfit criterion as an entry/removal criterion with a maximum effect number of 5. Consequently, using the outcome of the linear modelling analyses, we built the final model using manual multiple regression analyses and checked the residuals and the model quality data, and computed partial regression analyses including partial regression plots. All of the above analyses were performed using two-tailed tests, with an alpha level of 0.05 indicating statistical significance. We did not use false discovery rate (FDR) p corrections because all cytokines/chemokines/growth factors are associated in interconnected immune (cytokines + chemokines) and growth factor networks (Maes, Plaimas et al. 2021, Maes, Rachayon et al. 2022). Where needed, we employed transformations including logarithmic, square-root, and rank-based inversed normal (RINT) to normalize the data distribution of the indicators. We used IBM’s Windows version of SPSS 28 to perform all the above statistical analyses.

## Results

### Sociodemographic and clinical features of FE-MDMD

**Table 1** shows the sociodemographic and clinical data in FE-MDMD and healthy controls. There were no significant differences between patients and controls in age, sex, marital status, literate/illiterate ratio, and previous COVID-19 infection. BMI was somewhat lower in patients, while smoking was somewhat higher in patients. Accordingly, we have controlled all our analysis for BMI and smoking (and age, sex and previous COVID-19) by including these variables as covariates. The HAMD, HAMA, and SB sores were significantly higher in patients than in controls. There were no significant associations between COVID-19 and smoking and any of immune profiles or their single indicators. There were some effects of age and sex and, therefore, we controlled for these variables in regression analysis.

### Cytokines/chemokines/growth factors in FE-MDMD

**Figure 1** and ESF, Table 3, show that FE-MDMD patients have significantly increased levels of M1, M2, Th-1, Th-1/Th-2, neurotoxicity, IRS and CIRS as compared with healthy controls, while there were no significant differences in Th-2, Th-17, M1/M2, and IRS/CIRS. **Figure 2, Figure 3,** and ESF2, Tables 4 and 5, demonstrate that FE-MDMD patients show increased levels of sIL1RA, sIL2R, IL6, IL9, IL16, IL18, TNF-α, TNF-β, TRAIL, CCL2, CCL4, CCL5, CCL11, CCL27, CXCL1, CXCL9, CXCL10, MCSF, SCF, SCGF, SDF1, PDGFB, and HGF as compared with controls. There were no significant changes between patients and controls in IL-1α, IL-1β, IL2, IFN-γ, IL4, IL8, IL10, IL12p70, IL13, IL15, IL17, MIF1, CCL3, CCL7, LIF, G-CSF, GM-CSF, NGF, VEGF and FGF.

**Figure 1:**
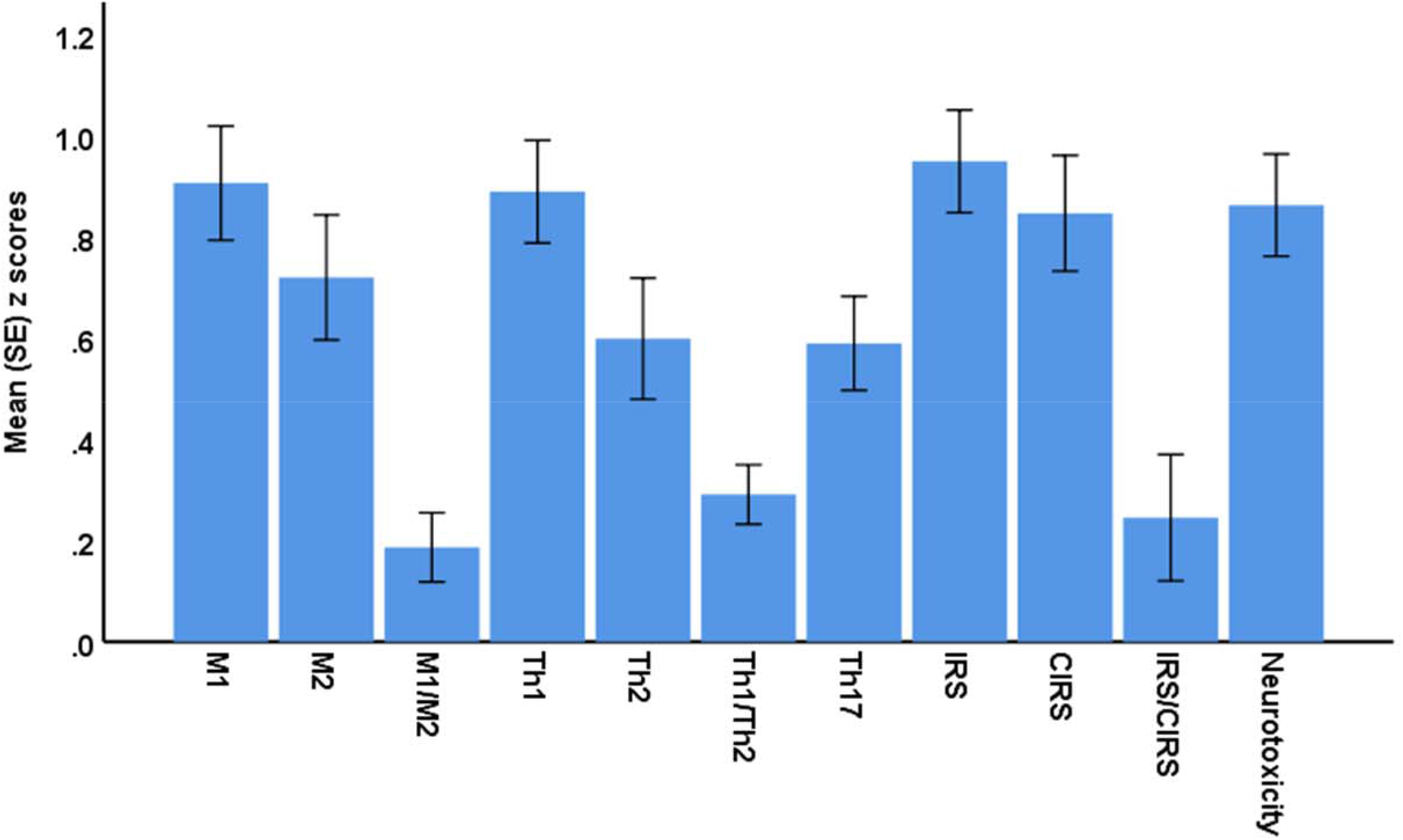
Mean values (standard error) of the z scores of immune profiles in patients with first-episode major dysmood disorder versus normal controls. The z scores of controls are set at zero. M1: classical M1, M2: alternative M2, Th: T helper, IRS: immune-inflammatory response system, CIRS: compensatory immunoregulatory system.

**Figure 2:**
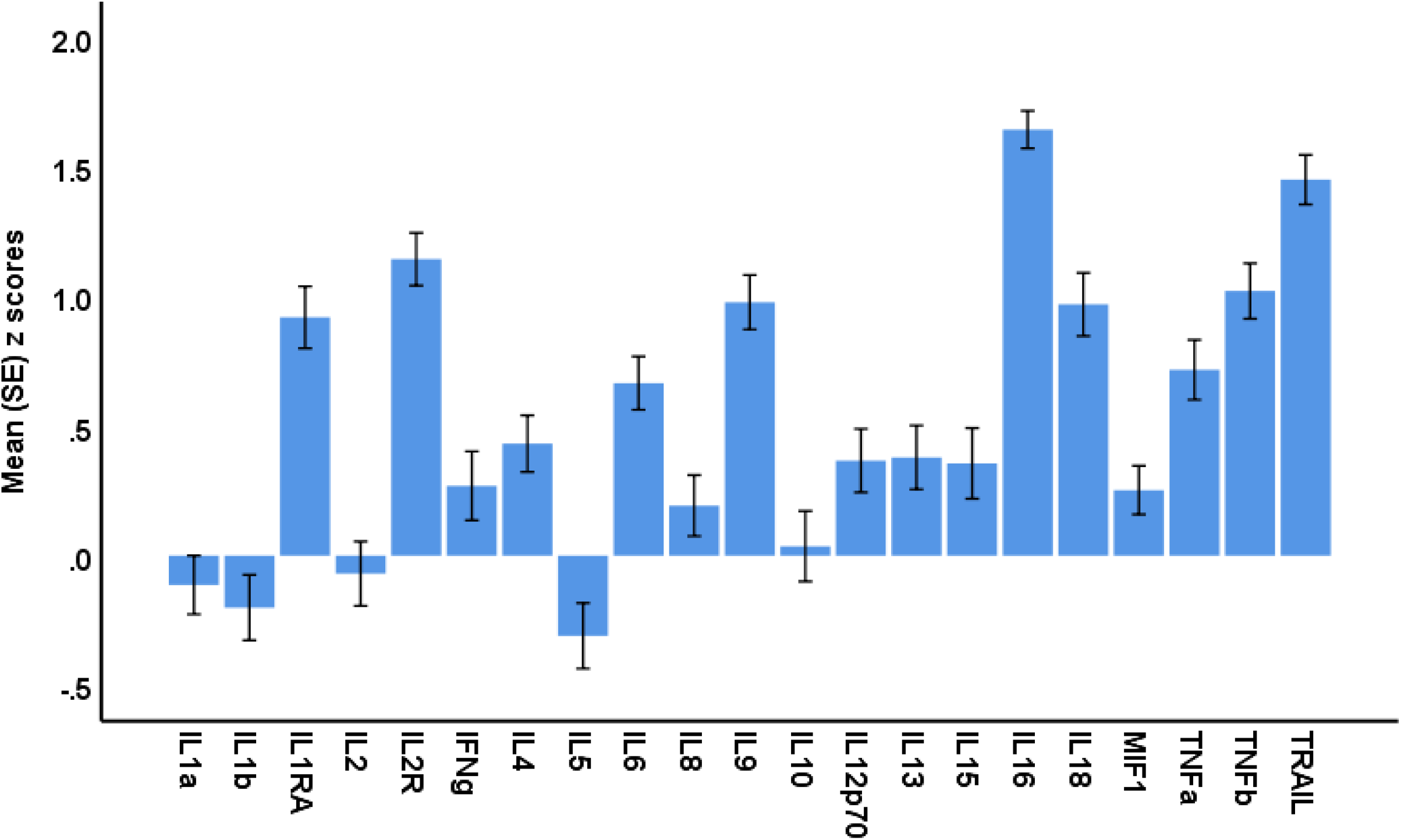
Mean values (standard error) of the z scores of cytokines in patients with first-episode major dysmood disorder versus normal controls. The z scores of controls are set at zero. IL: interleukin, IL1RA: interleukin receptor antagonist, MIF1: macrophage migration inhibitory factor-like protein, TNF: tumor necrosis factor, TRAIL: TNF-related apoptosis-inducing ligand.

**Figure 3:**
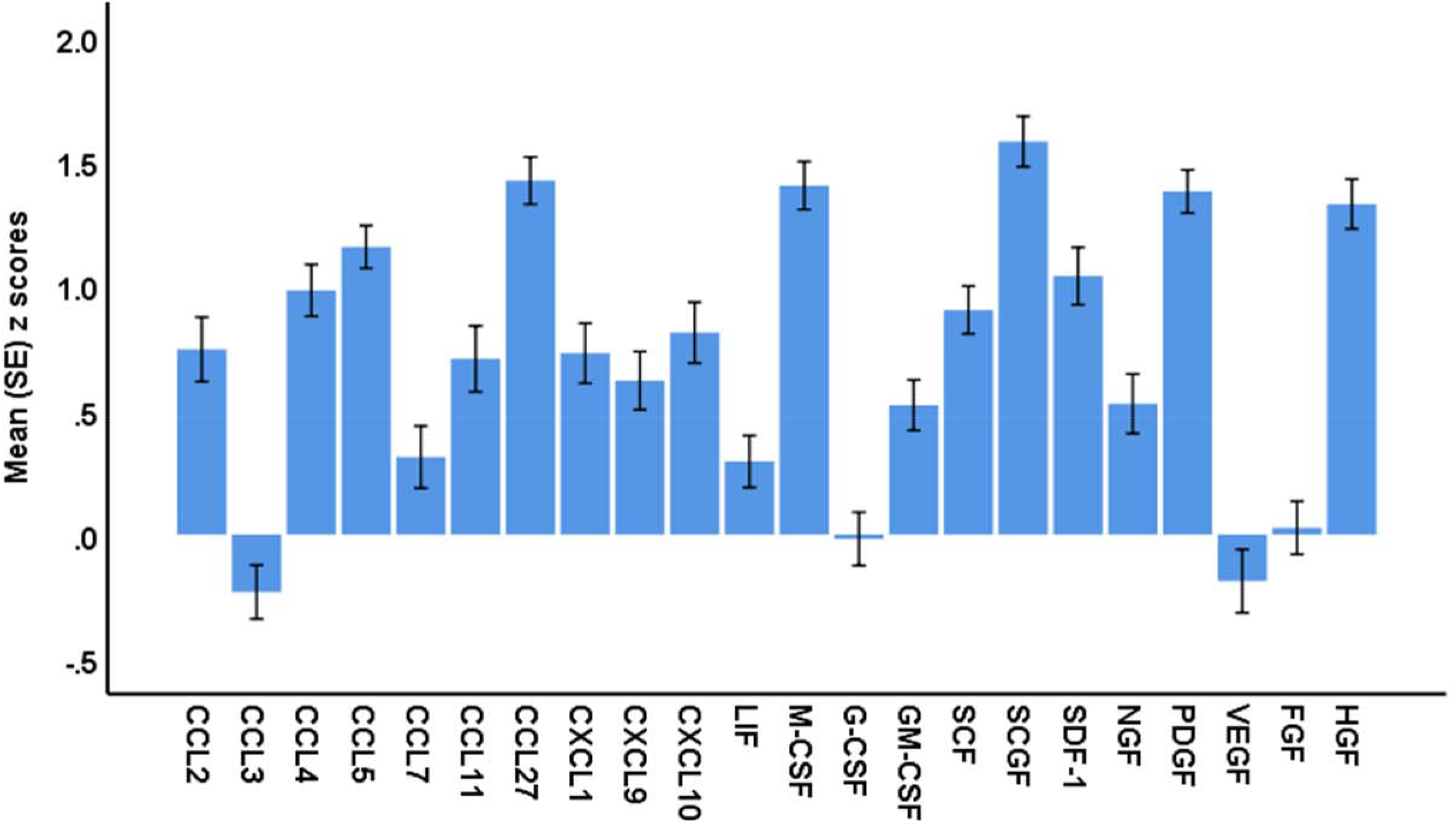
Mean values (standard error) of the z scores of chemokines/growth factors in patients with first-episode major dysmood disorder versus normal controls. The z scores of controls are set at zero. CC: C-C motif chemokine ligand, CXCL: C-X-C motif chemokine ligand, LIF: leukemia inhibitory factor, MCSF: macrophage colony-stimulating factor, GCSF: granulocyte CSF, GMCSF: granulocyte-macrophage CSF, SCF: stem cell factor, SCGF: stem cell growth factor-β, SDF1: stromal cell-derived factor-1, NGF: nerve growth factor, PDGFB: platelet-derived growth factor, VEGF: vascular endothelial growth factor, FGF: fibroblast growth factor, HGF: hepatocyte growth factor.

**Table 3:** Results of automatic linear modelling analyses (best subsets with overfit prevention criterion) with the Hamilton Depression Rating Scale (HAMD), Hamilton Anxiety Rating Scale (HAMA) and suicidal behaviors as dependent variables and cytokines, chemokines and growth factors as explanatory variables.

| Dependent Variables | Explanatory Variables | Parameter estimates |  |  |  | Model |  |  |  |
| --- | --- | --- | --- | --- | --- | --- | --- | --- | --- |
| | | $\beta$ | t | p | Importance | F | df | p | R2 |
| #1. HAMD | Model |  |  |  |  | 57.37 | 3/92 | <0.001 | 0.652 |
|  | IL-16 | 0.779 | 8.74 | <0.001 | 0.706 |  |  |  |  |
|  | sIL-1RA | -0.269 | -3.08 | 0.003 | 0.118 |  |  |  |  |
|  | PDGF | 0.369 | 3.69 | <0.001 | 0.093 |  |  |  |  |
| #1. HAMA | Model |  |  |  |  | 57.05 | 3/92 | <0.001 | 0.650 |
|  | IL-16 | 0.787 | 8.82 | <0.001 | 0.706 |  |  |  |  |
|  | sIL-1RA | -0.319 | -3.64 | <0.001 | 0.118 |  |  |  |  |
|  | PDGF | 0.312 | 3.97 | <0.001 | 0.093 |  |  |  |  |
| #3. Suicidal Behaviors | Model |  |  |  |  | 40.43 | 3/92 | <0.001 | 0.569 |
|  | IL-16 | 0.650 | 6.56 | <0.001 | 0.536 |  |  |  |  |
|  | PDGF | 0.427 | 4.89 | <0.001 | 0.298 |  |  |  |  |
|  | sIL-1RA | -0.355 | -3.65 | <0.001 | 0.166 |  |  |  |  |

IL: interleukin, sIL-1RA: soluble interleukin receptor antagonist, PDGF: platelet-derived growth factor, 95% CI: 95% confidence intervals.

### Intercorrelations

We employed Pearson’s correlations heatmaps to display the correlations between HAMD, HAMA, SB and the cytokines/chemokines/growth factors as shown in **Figures 4-6.** Figure 4 indicates that HAMA, HAMD and SB were significantly correlated with classical M1, but not M1/M2, Th-1, Th-1/Th-2, IRS, CIRS and neurotoxicity. Moreover, Th-2 and alternative M2 were correlated with HAMD and SB. Figure 5 shows that the clinical ratings were significantly correlated with IL-16, TRAIL, IL-18, TNF-α, TNF-β, IL-9, sIL-2R and sIL-1RA. Figure 6 shows that HAMA, HAMD and SB were significantly correlated with SCGF, PDGFB, M-CSF, CCL27, CCL5, HGF, CCL4, SDF1, SCF, CCL2, CXCL10, and CXCL9.

**Figure 4:**
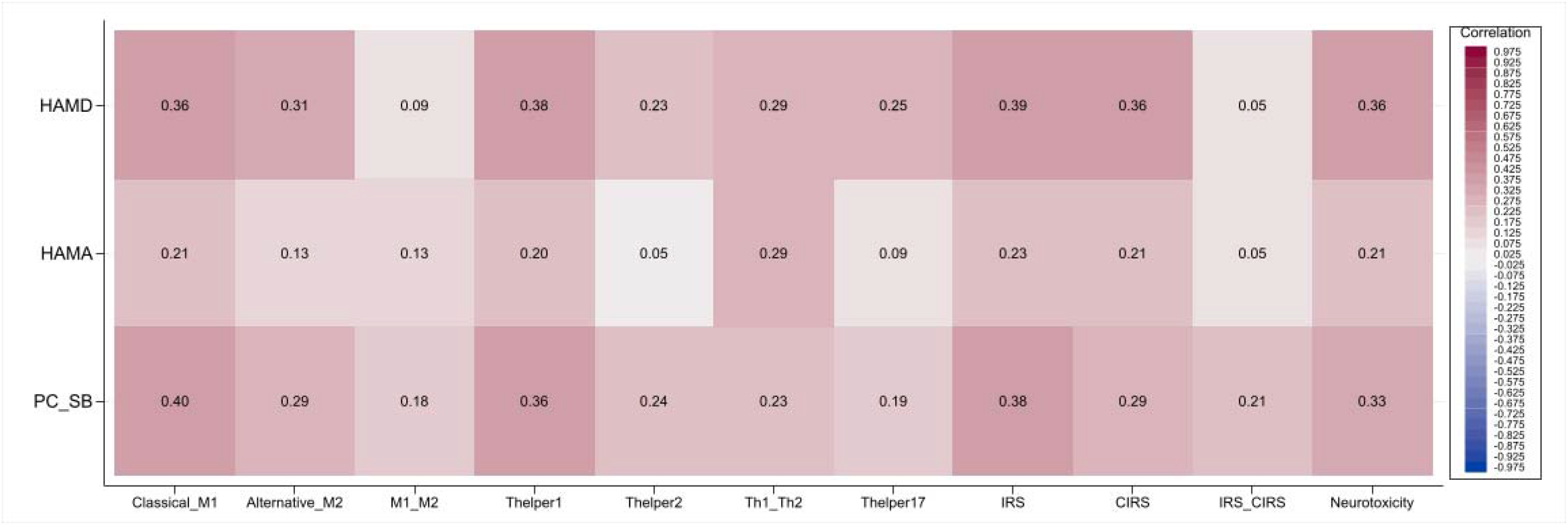
Pearson’s correlations heatmap. This correlogram displays the correlation between immune profiles and the Hamilton Depression Rating Scale (HAMD), Hamilton Anxiety Rating Scale (HAMA), and suicidal behaviors. Positive correlation coefficients are shaded in red, whereas negative correlation coefficients are shaded in blue. M1: classical M1, M2: alternative M2, Th: T helper, IRS: immune-inflammatory response system, CIRS: compensatory immunoregulatory system.

**Figure 5:**
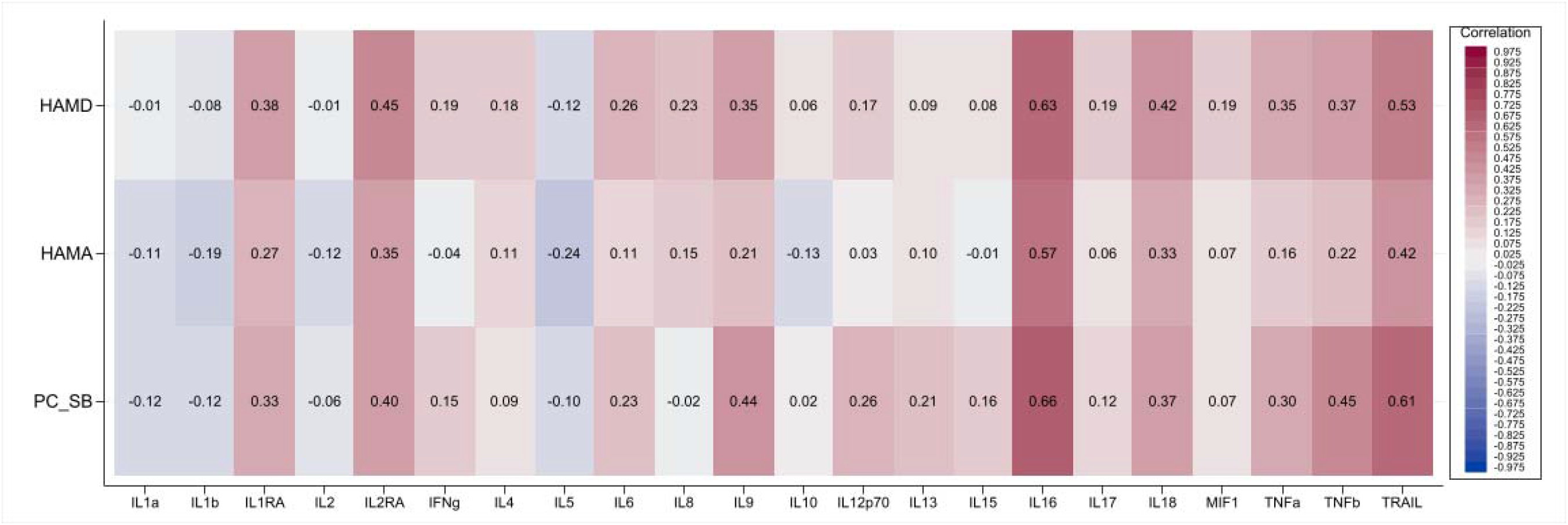
Pearson’s correlations heatmap. This correlogram displays the correlation between cytokines and the Hamilton Depression Rating Scale (HAMD), Hamilton Anxiety Rating Scale (HAMA), and suicidal behavior. Positive correlation coefficients are shaded in red, whereas negative correlation coefficients are shaded in blue. IL: interleukin, IL1RA: interleukin receptor antagonist, MIF1: macrophage migration inhibitory factor-like protein, TNF: tumor necrosis factor, TRAIL: TNF-related apoptosis-inducing ligand.

**Figure 6:**
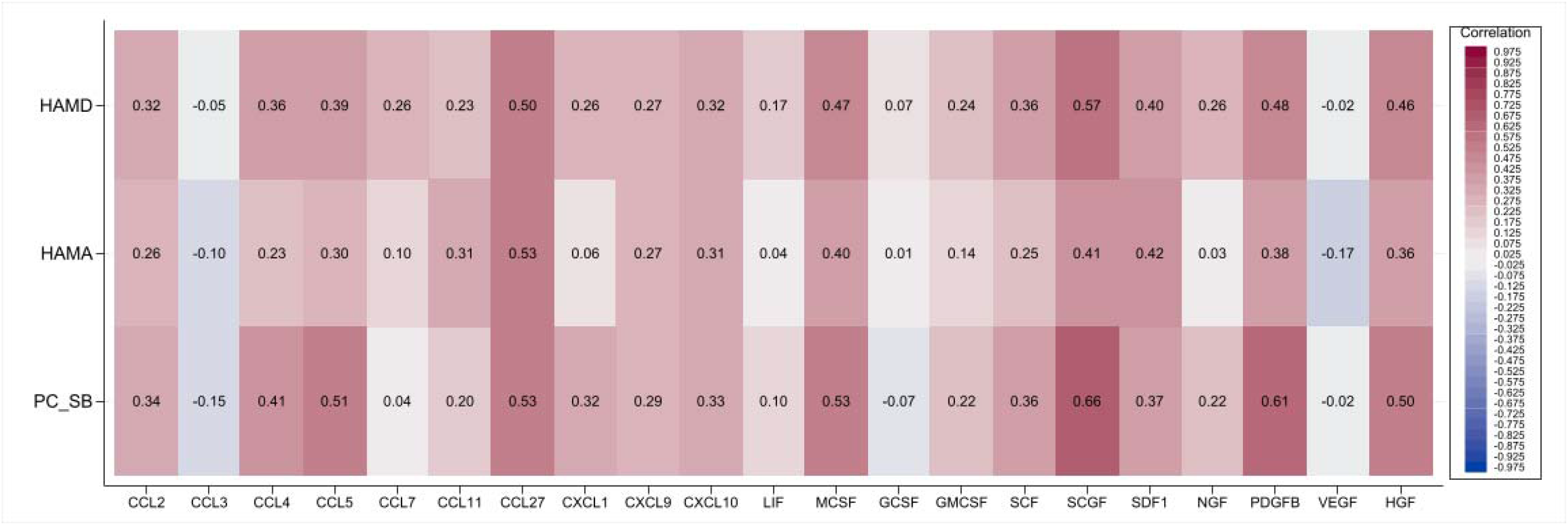
Pearson’s correlations heatmap. This correlogram displays the correlation between chemokines/growth factors and the Hamilton Depression Rating Scale (HAMD), Hamilton Anxiety Rating Scale (HAMA), and suicidal behaviors. Positive correlation coefficients are shaded in red, whereas negative correlation coefficients are shaded in blue. CC: C-C motif chemokine ligand, CXCL: C-X-C motif chemokine ligand, LIF: leukemia inhibitory factor, MCSF: macrophage colony-stimulating factor, GCSF: granulocyte CSF, GMCSF: granulocyte-macrophage CSF, SCF: stem cell factor, SCGF: stem cell growth factor-β, SDF1: stromal cell-derived factor-1, NGF: nerve growth factor, PDGFB: platelet-derived growth factor, VEGF: vascular endothelial growth factor, FGF: fibroblast growth factor, HGF: hepatocyte growth factor.

### Prediction of HAMA, HAMD and SB

**Table 2** shows the results of multiple regression analyses with HAMA, HAMD and SB as the dependent variables and the immune profiles as explanatory variables, while allowing for the effects of BMI, age, sex, smoking, and previous COVID-19 infection. Regression #1 illustrates that a significant proportion of the variance (25.6%) in the HAMD score was explained by IRS (positively associated), and Th-2 (inversely associated). Our findings in regression #2 indicate that 23.3% of the variance in the total HAMA score can be attributed to the positive relationship with IRS and a negative association with Th-2. SB was best predicted by M1 which explained 15.5% of the variance.

**Table 3** shows the results of automatic linear modelling analyses with overfit prevention that delineated the best subset of cytokines/chemokines/growth factors predicting the clinical scores. Again, we allowed for the effects of age, sex, BMI, smoking and COVID-19 by entering the variables as covariates, but none of these was significant. Table3, regression #1 indicates that IL-16, and PDGFB (all positively associated) and sIL-1RA (negatively associated) were the best predictors of HAMD explaining 65.2% of the variance. Regression #2 and #3 show that the same variables explained a large part of the variance in the total HAMA and SB scores.

### Cytokines/chemokines/growth factors and FE-MDMD

ESF, Table 6, displays that antidepressant-treated FE-MDMD patients compared to drug-naive FE-MDMD patients have significantly lower neurotoxicity, and CIRS scores, whereas the IRS/CIRS ratio was significantly higher in antidepressant-treated patients. Moreover, ESF, Tables 7 and 8 demonstrate significantly lower levels of IL-1α, sIL-2RA, IL4, IL8, IL17, CCL3, CCL7 and G-CSF in antidepressant-treated FE-MDMD patients as compared with controls.

## Discussion

### Immune Profiles in FE-MDMD

The first major finding of the current study is that FE-MDMD is characterized by activated IRS, CIRS, M1, M2, and Th-1 phenotypes, whereas no significant alterations in Th-2 and Th-17 were observed.

Importantly, we discovered that the Th-1/Th-2 ratio increased significantly, indicating Th-1 polarization during FE-MDMD. In stimulated whole blood cell cultures, the Th-1 profile was also elevated in patients with MDMD compared to controls and patients with SDMD (Maes, Rachayon et al. 2022). These findings provide additional evidence that T cell activation, as demonstrated by flow cytometry, is a key factor in severe MDD (Maes, Bosmans et al. 1990, Maes, Bosmans et al. 1991, Maes, Lambrechts et al. 1992, Maes, Stevens et al. 1993). Recent meta-analyses indicate that persons with MDD express more activated T cells and have a higher CD4/CD8 ratio (Foley, Parkinson et al. 2023, Sørensen, Borbye-Lorenzen et al. 2023).

Moreover, in the current study, there were no alterations in the M1/M2 and IRS/CIRS ratios. These findings are in agreement with our previous study showing activated IRS (M1 and Th-1) as well as CIRS in stimulated culture supernatants of MDMD patients (Maes, Rachayon et al. 2022). These findings indicate that a new equilibrium between enhanced M1/IRS and M2/CIRS profiles is established in FE-MDMD and that increased CIRS/M2 activity may restrain the activated IRS thereby establishing new homeostatic setpoints (Maes and Carvalho 2018).

### Cytokines/chemokines/growth factors in FE-MDMD

The second major finding of our study is that a) products of M1 macrophages (e.g. IL-6, TNF-α, CCL2, CXCL8, CXCL10, CXCL9) are considerably elevated, whereas IL-1β, IL-12p70, and IL-15 are not; b) M2 macrophage constituents such as PDGF and IL-1RA levels are increased, whereas IL-10, IL-4, and VEGF are not; c) Th-1 products such as IL-16, sIL-2R, TNF-α and TNF-β are significantly increased, whereas other important Th-1 players are not, including IFN-α2, IFN-γ, IL-2 and IL-12p70; d) IL-17, is not elevated in FE-MDMD; and e) some (sIL-1RA, sIL-2R, IL-5 and IL-9), but not all (IL-4, IL-10 and IL-13) CIRS products are altered in FE-MDMD.

Both the current study and Maes et al. (2022) established overlapping elevations of sIL-1RA, IL-5, IL-9, CXCL-10, PDGF, CCL5, and TNF-α levels in MDMD. In contrast to Maes et al. (2022), the present study was unable to detect increases in IL-15, FGF, IFN-γ, and VEGF in MDMD. The latter analytes are, however, extremely difficult to quantify in serum but are easily quantifiable in culture supernatant (Maes, Rachayon et al. 2022).

A recent meta-analysis revealed that first-episode MDD is associated with elevated levels of IL-6, TNF-α, IL-1β, IL-2, and C-reactive protein, while IL-8 levels are reduced (Çakici, Sutterland et al. 2020). Some studies found significant increases in IL-6, TNF-α, IL-1β, IL-12, IL-17, IL-9, IL-10, and IL-5 in FED (Jiang, Zhang et al. 2017, Çakici, Sutterland et al. 2020, Kakeda, Watanabe et al. 2020, Lan, Zhou et al. 2021). Lan et al., found that 16 out of 19 distinct cytokines significantly differed between FED and healthy controls, including GM-CSF, fractalkine, IFN-γ, IL-10, MIP-3α, IL-12p70, IL-13, IL-17A, IL-1β, IL-2, IL-23, IL-5, IL-6, and TNF-α (Lan, Zhou et al. 2021). As in our study, previous reports could not establish differences in IL-4 and IL-10 between FED and healthy controls (Lan, Zhou et al. 2021, Yang, Liu et al. 2021). However, in cerebrospinal fluid (CSF), there was no difference in 25 cytokines/chemokines between individuals with recent-onset depression and healthy controls (Sørensen, Borbye-Lorenzen et al. 2023). Other studies were unable to detect alterations in CSF levels of TNF-α, TNF-β, sIL-1RA, and sIL-2R in FED (Yang, Liu et al. 2021). Lee et al. (Lee, Song et al. 2020) reported that adolescent patients with FED had decreased levels of IL-2, IFN-γ, TNF-α, and IL-10.

These contradictory outcomes may be attributable to differences in the media employed to assay the immune mediators (e.g., serum versus CSF). Importantly, there are two phenotypes of MDD (see Introduction), and failure to account for the biomarker differences between MDMD and SDMD results in erroneous conclusions (Maes, Moraes et al. 2021, Simeonova, Stoyanov et al. 2021, Maes 2022, Maes, Rachayon et al. 2022, Maes, Vasupanrajit et al. 2023). In addition, using the MDD diagnosis prevents the discovery of pathways due to the following issues: a) low reliability and validity of the MDD diagnosis; b) people with normal responses of sadness, feeling blue, grieving, and demoralisation are often lumped together with MDD because the criteria do not allow a sharp demarcation; and c) patients with an acute phase, partial remission, and remission are often lumped together (Maes and Almulla 2022, Maes, Rachayon et al. 2022). Moreover, our recent meta-analyses revealed that a large number of individuals do not know how to measure and statistically analyse cytokine levels (e.g., how to censor and manage values below the detection limit) (Vasupanrajit, Jirakran et al. 2021, Vasupanrajit, Jirakran et al. 2022).

Last but not least, one wonders why researchers use an unreliable binary diagnosis (MDD) when it is more appropriate to use quantitative phenome scores computed using machine learning techniques (Maes and Almulla 2022, Maes, Rachayon et al. 2022). Examining the associations between immune profiles and their single indicators and quantitatively assessing the phenome of depression is the most essential method (Maes and Almulla 2022, Maes, Rachayon et al. 2022), as discussed in the next section.

### Associations among immune profiles and severity of depression and anxiety

The third major finding of the current study is that a) a significant portion of the variance in the severity of depression (25.6%) and anxiety (23.3%) could be predicted by IRS (positive) and Th-2 (inverse) profiles; and b) IL-16 is, by far, the most important cytokine associated with the depression phenome, followed by TRAIL, sIL-2R, IL-18, CCL27, M-CSF, SDF-1, and HGF, and then at a distance by TNF-α and TNF-β, sIL-1RA, IL-9, SCF, and SCGF.

The significance of Th-1 polarisation in FE-MDMD is further supported by the observation that the Th-2 immune profile, but not the Treg profile, is negatively associated with the severity of depression and anxiety. As a result, relative decreases in Th-2 functions may impact the negative immunoregulatory effects of Th-2, resulting in greater effects of Th-1 and IRS activities. The IRS-CIRS theory of depression describes such relative deficits in CIRS activities in depression (Maes, Rachayon et al. 2022).

IL-16 is most likely a crucial component of FE-MDMD that may trigger T cell activation. IL-16 signaling does, in fact, take place through the CD4 molecule on Th cells, activating CD4+ cells, upregulating CD25+ and HLA-DR+ markers, and initiating the IRS (Mathy, Scheuer et al. 2000, Hall, Cullen et al. 2016). These results emphasize T-cell activation’s significance in severe MDD/MDMD even more (Maes, Bosmans et al. 1990, Maes, Lambrechts et al. 1992, Maes, Stevens et al. 1993). According to previous research, IL-16 was significantly correlated with current depression (Timothy, Helena et al. 2018). Proinflammatory cytokines, including IL-16, were highly associated with depression and neuroticism in hepatitis C patients six months after the start of cytokine-based treatment (Pawlowski, Radkowski et al. 2014). In animal models, prenatal depression was linked to higher levels of cytokines, including IL-16, in the hippocampus and prefrontal cortex immediately after birth (Posillico and Schwarz 2016, Haim, Julian et al. 2017). Intriguingly, IL-16 has been shown to influence neuroinflammation in animal models of autoimmune encephalomyelitis by attracting and activating CD4+ immune cells (Hridi, Barbour et al. 2021).

The second most important mediator in FE-MDMD that positively mediate the inflammatory response, apoptosis, and glial cell proliferation is elevated TNF signaling as indicated by increased levels of TRAIL, TNF-α, and TNF-β (String, functional protein association networks (string-db.org)).

It is important to stress that the activated cytokines/chemokines/growth factors are strongly interconnected and act within a tight protein-protein interaction (PPI) network (Maes, Rachayon et al. 2022). Also, the cytokines/chemokines/growth factors that are elevated in our FE-MDMD patients form a tight network as can be observed using STRING (String, <u>STRING: functional protein association networks (string-db.org).</u> This tight network shows an adequate average clustering coefficient of 0.772 and an average node degree of 9.47 (PPI enrichment value < 10^-16^). The subnetwork of the growth factors that are elevated in FE-MDMD indicate positive regulation of cell migration and immune cell population proliferation (String, <u>STRING: functional protein association networks (string-db.org).</u> Previously, it was detected that growth factor abnormalities are part of the same PPI network as pro-inflammatory cytokines, indicating that they play a crucial role in the development of FE-MDMD (Maes, Rachayon et al. 2022).

### Associations among immune profiles and suicidal behaviors

Another major finding is that suicidal behavior in FE-MDMD is associated with increased M1 activation and that IL-16 and PDGFB (positively) and lowered sIL-1RA explain 56.9% of the variance in suicidal behaviors. This indicates that activated M1, Th-1 (IL-16) and the growth factor subnetwork (PDGFB) are major pathways leading to suicidal behavior in FE-MDMD. Prior research demonstrated that elevated IRS, neuroimmunotoxicity, and growth factor profiles predict a substantial portion of the depression phenome, including suicidal behaviors (Maes, Rachayon et al. 2022). Recent meta-analyses demonstrating that IRS pathways contribute to suicidal behavior, are consistent with these findings (Vasupanrajit, Jirakran et al. 2021, Vasupanrajit, Jirakran et al. 2022).

### Increased neuroimmunotoxicity in FE-MDMD

Perhaps the most relevant finding is that the FE-MDMD phenome features are strongly associated with a neurotoxicity profile consisting of serum IL-6, TNF-α, TRAIL, IL-16, CCL2, CCL11, CXCL1, CXCL10, and M-CSF. As such, increased Th-1 cell (IL-16) activation and, secondarily, M1 cell activation appear to be the primary drivers of neuro-affective toxicity in MDMD. Using culture supernatant from stimulated whole blood, we determined that CCL5, CXCL10, TNF-α, CXCL8, IL-17, and IFN-γ were the neurotoxic substances that were elevated in MDMD. The neurotoxicity theory of MDD postulates that neurotoxic cytokines and chemokines exert neuro-affective toxicity, culminating in depression through injury to astroglial and neuronal projections (Maes and Carvalho 2018, Al-Hakeim, Al-Naqeeb et al. 2023).

### Effects of antidepressants on immune-inflammatory pathways in FE-MDMD

The fifth major finding of this study is that a) drug-naive FE-MDMD patients displayed significantly increased neurotoxicity, Th-17, and CIRS, and a decreased IRS/CIRS ratio compared to patients who took antidepressants for at least three weeks; and b) drug-naive patients exhibit significantly increased production of IL-1β, IL-2RA, IL-4, CXL8, CCL3, CCL7, and G-CSF. As a result, antidepressants have contradictory effects on the immune system, diminishing both inflammatory mediators (Th-17, IL-1β, CXL8, CCL3, CCL7, and G-CSF) and CIRS functions (CIRS, sIL-2R, and IL-4). In addition, the increase in the IRS/CIRS ratio indicates that antidepressant treatment does not normalize the IRS and neurotoxicity responses and, actually impacts IRS activation by decreasing CIRS functions. Therefore, the anti-inflammatory and immunoregulatory effects of antidepressants were probably overemphasised in the past (Nässberger and TräskmanLBendz 1993, Maes, Song et al. 1999, Roumestan, Michel et al. 2007, Tynan, Weidenhofer et al. 2012, Lan, Zhou et al. 2021).

## Limitations

Our results would be more interesting if we could evaluate oxidative and nitrosative stress toxicity in FE-MDMD patients since these pathways are strongly intertwined with IRS activation (Moylan, Berk et al. 2014). Another potential limitation is the possible impact of COVID-19. However, no patients with acute COVID-19 were included, nor did any subjects have Long COVID. In addition, the COVID patients included in this study had previously experienced mild COVID-19 more than 6 months prior to recruitment. The depression and anxiety symptoms due to Long COVID are strongly predicted by severe and critical COVID-19, while mild infection is not really associated with later Long COVID symptoms (Al-Hakeim, Al-Rubaye et al. 2022). As such, we limited the possible impact of previous COVID-19 on our results. Furthermore, statistically accounting for the potential effects of previous mild COVID-19 revealed that this had no significant impact on the results. The major strength of the current study is that we recruited FE-MDMD, including drug-naïve FE-MDMD, patients.

## Conclusions

The present study presents substantial evidence of activated immune-inflammatory responses, Th-1 polarization and increased immune-related neurotoxicity in FE-MDMD, including drug-naïve patients. Thus, we found activated M1, M2, and Th-1 phenotypes along with upregulated IRS, CIRS and neurotoxicity pathways. Upregulation of IL-16 is the key cytokine associated with FE-MDMD and, therefore, IL-16 or CD4 activation are probably the most promising novel drug targets to treat FE-MDMD. Moreover, our results emphasize that antidepressants have mixed effects on the immune system, namely anti-IRS and anti-CIRS effects.

## Data Availability

The dataset generated during and/or analyzed during the current study will be available from the corresponding author (MM) upon reasonable request and once the dataset has been fully exploited by the authors.

## Author’s contributions

All authors contributed to the manuscript and read and approved the final draft.

## Institutional Review Board Statement

The College of Medical Technology at The Islamic University of Najaf, Iraq (18/2021) approved the research project. Our IRB follows the International Guideline for Human Research Safety, as well as the World Medical Association Declaration of Helsinki, The Belmont Report, the CIOMS Guideline, and the International Conference on Harmonization of Good Clinical Practice, and our study was conducted in accordance with all applicable Iraqi and international ethics and privacy laws. (ICH-GCP).

## Informed Consent Statement

All the controls and patients or their parents/legal guardians provided written signed consent.

## Acknowledgements

The authors acknowledge the Al-Hakeem General Hospital workers for their efforts in data collection.

