## supplementary file for "T helper-1 activation via interleukin-16 is a key phenomenon in the acute phase of severe, first-episode major depressive disorder and suicidal behaviors"

**ELECTRONIC SUPPLEMENTARY FILE (ESF)**

**ESF, Table 1**. Overview of the cytokines, chemokines, and growth factors measured in the current study

| **Protein abbreviations** | **Gene Symbol** | **> OOR (%)** | **Protein name / alias** |
| --- | --- | --- | --- |
| **IFN-α2** | **IFNA2** | 7.0 | Interferon-α2 |
| **IFN-γ** | **IFNG** | 100 | Interferon-γ |
| **IL-1α** | **IL1A** | 100 | Interleukin-1α |
| **IL-1β** | **IL1B** | 80.7 | Interleukin-1β |
| **sIL-1RA** | **IL1RN** | 79.8 | Soluble interleukin-1 receptor antagonist |
| **IL-2** | **IL2** | 56.5 | Interleukin-2 |
| **IL-2R** | **IL2RA** | 100.0 | Soluble interleukin-2 receptor |
| **IL-3** | **IL3** | 8.8 | Interleukin-3 |
| **IL-4** | **IL4** | 100 | Interleukin-4 |
| **IL-5** | **IL5** | 44.7 | Interleukin-5 |
| **IL-6** | **IL6** | 79.8 | Interleukin-6 |
| **IL-7** | **IL7** | 7.9 | Interleukin-7 |
| **IL-9** | **IL9** | 100 | Interleukin-9 |
| **IL-10** | **IL10** | 67.5 | Interleukin-10 |
| **IL-12p70** | **IL12RB1** | 52.6 | Interleukin-12 p70 |
| **IL-12p40** | **IL12RB1** | 6.1 | Interleukin-12 p40 |
| **IL-13** | **IL13** | 93.0 | Interleukin-13 |
| **IL-15** | **IL15** | 72.8 | Interleukin-15 |
| **IL-16** | **IL16** | 100 | Interleukin-16 |
| **IL-17** | **IL17A** | 28.9 | Interleukin-17 |
| **IL-18** | **IL18** | 100 | Interleukin-18 |
| **TNF-α** | **TNF** | 100 | Tumor necrosis factor-α |
| **TNF-β** | **LTA** | 100 | Tumor necrosis factor-β or lymphotoxin-alpha (LT-α) |
| **TRAIL** | **TNFSF10** | 100 | TNF-related apoptosis-inducing ligand (TRAIL) or tumor necrosis factor ligand superfamily member 10 (TNFSF10) |
| **LIF** | **LIF** | 92.1 | Leukemia inhibitory factor or IL-6 famiy cutokine |
| **MIF** | **MIF** | 100 | Macrophage migration inhibitory factor-like protein (MIF) or glycosylation-inhibiting factor |
| **G-CSF** | **CSF3** | 100 | Granulocyte colony stimulating factor (G-CSF) or colony stimulating factor 3 (CSF3) |
| **M-CSF** | **CSF1** | 100 | Macrophage colony-stimulating factor (M-CSF) or colony stimulating factor 1 (CSF1) |
| **GM-CSF** | **CSF2** | 86.8 | Granulocyte-macrophage colony-stimulating factor (GM-CSF) or colony-stimulating factor 2 (CSF2) |
| **CCL2 or MCP1** | **CCL2** | 100 | C-C motif chemokine ligand 2 (CCL2) or monocyte chemoattractant protein 1 (MCP1) |
| **CCL3 or MIP-1α** | **CCL3** | 93 | C-C motif Chemokine ligand 3 (CCL3) or macrophage inflammatory protein 1-alpha (MIP-1α) |
| **CCL4 or MIP-1β** | **CCL4** | 100 | C-C motif chemokine ligand 4 (CCL4) or macrophage inflammatory protein 1β (MIP-1β) or lymphocyte activation gene 1 protein |
| **CCL5 or RANTES** | **CCL5** | 66.7 | C-C motif chemokine ligand 5 (CCL5) or regulated upon activation, normally T-expressed, and presumably Secreted (RANTES) |
| **CCL7 or MCP3** | **CCL7** | 75.4 | C-C motif chemokine ligand 7 (CCL7) or monocyte-chemotactic protein 3 (MCP3). |
| **CCL11 or Eotaxin** | **CCL11** | 100 | C-C motif chemokine ligand 11 (CCL11) or eosinophil chemotactic protein |
| **CCL27 or CTACK** | **CCL27** | 100 | C-C motif chemokine ligand 27 (CCL27) or cutaneous T-cell attracting chemokine (CTACK) |
| **CXCL1 or GRO-α** | **CXCL1** | 100 | C-X-C motif chemokine 1 (CXCL1) or growth-regulated alpha protein (GRO) |
| **CXCL8 or IL-8** | **CXCL8** | 100 | C-X-C motif chemokine ligand 8 (CXCL8) or interleukin-8 (IL-8) |
| **CXCL9 or MIG** | **CXCL9** | 100 | C-X-C motif chemokine ligand 9 (CXCL9) or monokine induced by gamma interferon (MIG) |
| **CXCL10 or IP10** | **CXCL10** | 100 | C-X-C motif chemokine ligand 10 (CXCL10) or Interferon gamma-induced protein 10 (IP10) |
| **CXCL12 or SDF-1α** | **CXCL12** | 100 | C-X-C motif chemokine 12 (CXCL12) or stromal cell-derived factor 1 (SDF-1α) |
| **FGF** | **FGF2** | 43.0 | Fibroblast growth factor 2 (FGF) or basic fibroblast growth factor |
| **HGF or SF** | **HGF** | 100 | Hepatocyte growth factor (HGF) or scatter factor (SF) |
| **BNGF** | **NGF** | 100 | β-nerve growth factor (NGF) |
| **PDGF** | **PDGFA** | 100 | Platelet derived growth factor (PDGF) |
| **SCF** | **KITLG** | 100 | Stem cell factor (SCF) or Kit ligand (KITLG) |
| **SDF-1** | **SDF1** | 100 | Stromal cell-derived factor |
| **SCGF-β or CLEC11A** | **CLEC11A** | 100 | Stem cell growth factor (SCGF) or C-type lectin domain family 11 member A (CLEC11A) |
| **VEGF** | **VEGFA** | 100 | Vascular endothelial growth factor (VEGF) |

Adapted from: *Maes M, Rachayon M, Jirakran K, Sodsai P, Klinchanhom S, Gałecki P, Sughondhabirom A, Basta-Kaim A. The Immune Profile of Major Dysmood Disorder: Proof of Concept and Mechanism Using the Precision Nomothetic Psychiatry Approach. Cells. 2022 Mar 31;11(7):1183. doi: 10.3390/cells11071183. PMID: 35406747; PMCID: PMC8997660.*

*Kalayasiri R, Dadwat K, Supaksorn T, ,Sirivichayakul S, Maes M. Methamphetamine (MA) use, MA dependence, and MA-induced psychosis are associated with increasing aberrations in the compensatory immunoregulatory system and interleukin-1α and CCL5 levels. medRxiv 2023.03.26.23287766; doi: https://doi.org/10.1101/2023.03.26.23287766*

**ESF, Table 2**. Description of the immune profiles used in this study

| **Immune Profile** | **Members** |
| --- | --- |
| **M1 macrophage** | IL-1β, IL-6, TNF-α, IL-12p70, IL-15, CCL2, CCL5, CXCL1, CXCL8, CXCL9, CXCL10 |
| **M2 macrophages** | IL-10, IL-4, IL-13, VEGF, PDGF, sIL-1RA |
| **z M1 – z M2** | zM1 – zM2 |
| **T helper (Th)-1** | IL-2, sIL-2R, IFN-a, IFN-γ, IL-12p70, IL-16, TNF-α, TNF-β |
| **Th-2** | IL-4, IL-5, IL-9, IL-13, IL-10, IL-6 |
| **z Th- z Th-2** | zTh-1 – zTh-2 |
| **Th-17** | IL-6, IL-17 |
| **IRS** | IL-1α, IL-1β, IL-6, TNF-α, IL-12p70, IL-15, IL-16, IL-17, IL-18, CCL2, CCL3, CCL4, CCL5, CCL7, CCL11, CXCL1, CXCL8, CXCL9, CXCL10, IL-2, IFN-α, IFN-γ, TNF-α, TNF-β, TRAIL, GM-CSF, M-CSF, G-CSF, SCGF |
| **CIRS** | IL-4, IL-10, sIL-1RA, sIL-2R |
| **z IRS – z CIRS** | Z IRS – z CIRS |
| **Neurotoxicity** | IL-1β, IL-6, TNF-α, TRAIL, IL-2, IFN-γ, IL-12p70, IL-16, IL-17, CCL2, CCL3, CCL5, CCL11, CXCL1, CXCL8, CXCL10, GM-CSF, M-CSF |

IRS: immune-inflammatory response system; CIRS: compensatory immunoregulatory system

Adapted from:

*Maes M, Rachayon M, Jirakran K, Sodsai P, Klinchanhom S, Gałecki P, Sughondhabirom A, Basta-Kaim A. The Immune Profile of Major Dysmood Disorder: Proof of Concept and Mechanism Using the Precision Nomothetic Psychiatry Approach. Cells. 2022 Mar 31;11(7):1183. doi: 10.3390/cells11071183. PMID: 35406747; PMCID: PMC8997660.*

*Kalayasiri R, Dadwat K, Supaksorn T, ,Sirivichayakul S, Maes M. Methamphetamine (MA) use, MA dependence, and MA-induced psychosis are associated with increasing aberrations in the compensatory immunoregulatory system and interleukin-1α and CCL5 levels. medRxiv 2023.03.26.23287766; doi: https://doi.org/10.1101/2023.03.26.23287766*

**ESF, Table 3**. Differences in immune cells and immune profiles between first-episode major dysmood disorder (FE-MDMD) and healthy controls (HC).

| **Variables (z scores)** | **HC (n=40)** | **FE-MDMD (n=71)** | **F/X^2^** | **df** | **p-value** |
| --- | --- | --- | --- | --- | --- |
| Classical M1 | 0.145(0.171) | 0.955(0.112) | 15.666 | 1/89 | <0.001 |
| Alternative M2 | 0.088(0.183) | 0.787(0.120) | 10.187 | 1/89 | 0.002 |
| z M1 – z M2 | 0.057(0.122) | 0.168(0.079) | 0.586 | 1/89 | 0.446 |
| Th-1 | 0.204(0.169) | 0.947(0.110) | 13.519 | 1/89 | <0.001 |
| Th-2 | 0.237(0.177) | 0.633(0.115) | 3.523 | 1/89 | 0.064 |
| z Th-1 – z Th-2 | -0.033(0.103) | 0.314(0.067) | 7.923 | 1/89 | 0.006 |
| Th-17 | 0.216(0.184) | 0.617(0.120) | 3.347 | 1/89 | 0.071 |
| IRS | 0.170(0.170) | 0.999(0.111) | 16.747 | 1/89 | <0.001 |
| CIRS | 0.075(0.182) | 0.886(0.119) | 13.965 | 1/89 | <0.001 |
| z IRS – z CIRS | 0.225(0.196) | 0.271(0.128) | 0.039 | 1/89 | 0.844 |
| Neurotoxicity | 0.177(0.175) | 0.900(0.114) | 11.980 | 1/89 | <0.001 |

M: macrophage, Th: T helper, IRS: immune-inflammatory response system, CIRS: compensatory immunoregulatory response system

All results of GLM analysis with age, sex, body mass index, smoking, and previous mild-moderate COVID-19 (entered as dummy) as covariates

**ESF, Table 4**. Differences in cytokines between first-episode major dysmood disorder (FE-MDMD) and healthy controls (HC).

| **Variables (z scores)** | **HC**  **(n=40)** | **FE-MDMD (n=71)** | **F/X2** | **df** | **p-value** |
| --- | --- | --- | --- | --- | --- |
| IL-1α | 0.230(0.201) | -0.066(0.131) | 1.522 | 1/89 | 0.221 |
| IL-1β | 0.203(0.198) | -0.233(0.129) | 3.409 | 1/89 | 0.068 |
| IL-1RA | -0.113(0.184) | 0.980(0.120) | 24.778 | 1/89 | <0.001 |
| IL-2 | 0.206(0.195) | -0.006(0.127) | 0.828 | 1/89 | 0.365 |
| IL-2R | 0.089(0.174) | 1.154(0.113) | 26.385 | 1/89 | <0.001 |
| IFN-γ | 0.120(0.197) | 0.334(0.129) | 0.831 | 1/89 | 0.364 |
| IL-4 | 0.137(0.205) | 0.439(0.134) | 1.530 | 1/89 | 0.219 |
| IL-5 | 0.143(0.189) | -0.308(0.124) | 3.990 | 1/89 | 0.049 |
| IL-6 | 0.196(0.193) | 0.690(0.126) | 4.609 | 1/89 | 0.035 |
| IL-8 | -0.088(0.191) | 0.100(0.125) | 0.677 | 1/89 | 0.413 |
| IL-9 | 0.127(0.160) | 1.077(0.104) | 24.744 | 1/89 | <0.001 |
| IL-10 | 0.113(0.188) | 0.078(0.123) | 0.024 | 1/89 | 0.877 |
| IL-12p70 | 0.166(.179) | 0.402(0.117) | 1.218 | 1/89 | 0.273 |
| IL-13 | 0.149(0.192) | 0.333(0.125) | 0.641 | 1/89 | 0.426 |
| IL-15 | 0.208(0.191) | 0.460(0.125) | 1.220 | 1/89 | 0.272 |
| IL-16 | 0.011(0.123) | 1.652(0.080) | 125.954 | 1/89 | <0.001 |
| IL-18 | -0.069(0.186) | 0.984(0.121) | 22.489 | 1/89 | <0.001 |
| MIF-1 | -0.092(0.193) | 0.129(0.126) | 0.915 | 1/89 | 0.341 |
| TNF-α | 0.179(0.182) | 0.767(0.119) | 7.287 | 1/89 | 0.008 |
| TNF-β | 0.125(0.165) | 1.131(0.108) | 25.966 | 1/89 | <0.001 |
| TRAIL | 0.039(0.147) | 1.496(0.096) | 68.735 | 1/89 | <0.001 |

IL: interleukin, IFN: interferon, IL1RA: interleukin receptor antagonist, MIF1: macrophage migration inhibitory factor-like protein, TNF: tumor necrosis factor, TRAIL: TNF-related apoptosis-inducing ligand.

All results of GLM analysis with age, sex, body mass index, smoking, and previous mild COVID-19 (entered as dummy) as covariates

**ESF, Table 5**. Differences in chemokines and growth factors between first-episode major dysmood disorder (FE-MDMD) and healthy controls (HC).

| **Variables (z scores)** | **HC**  **(n=40)** | **FE-MDMD**  **(n=71)** | **F/X2** | **df** | **p-value** |
| --- | --- | --- | --- | --- | --- |
| CCL2 | -0.048(0.183) | 0.802(0.119) | 15.166 | 1/89 | <0.001 |
| CCL3 | 0.197(0.200) | -0.228(0.131) | 3.157 | 1/89 | 0.079 |
| CCL4 | 0.165(0.178) | 1.038(0.116) | 16.957 | 1/89 | <0.001 |
| CCL5 | 0.116(0.164) | 1.276(0.107) | 35.101 | 1/89 | <0.001 |
| CCL7 | 0.070(0.191) | 0.317(0.124) | 1.174 | 1/89 | 0.281 |
| CCL11 | -0.002(0.189) | 0.655(0.123) | 8.507 | 1/89 | 0.004 |
| CCL27 | 0.055(0.153) | 1.365(0.100) | 51.541 | 1/89 | <0.001 |
| CXCL1 | 0.152(0.161) | 0.856(0.105) | 13.351 | 1/89 | <0.001 |
| CXCL9 | -0.105(0.176) | 0.589(0.115) | 10.839 | 1/89 | 0.001 |
| CXCL10 | -0.078(0.192) | 0.766(0.125) | 13.608 | 1/89 | <0.001 |
| LIF | 0.129(0.197) | 0.365(0.128) | 1.009 | 1/89 | 0.318 |
| M-CSF | 0.082(0.158) | 1.412(0.103) | 49.862 | 1/89 | <0.001 |
| G-CSF | 0.105(0.208) | -0.031(0.136) | 0.300 | 1/89 | 0.585 |
| GM-CSF | 0.211(0.183) | 0.573(0.119) | 2.763 | 1/89 | 0.100 |
| SCF | 0.134(0.175) | 0.934(0.114) | 14.640 | 1/89 | <0.001 |
| SCGF | -0.026(0.138) | 1.559(0.090) | 92.633 | 1/89 | <0.001 |
| SDF-1 | 0.019(0.170) | 0.967(0.111) | 21.822 | 1/89 | <0.001 |
| NGF | 0.255(0.171) | 0.631(0.112) | 3.374 | 1/89 | 0.070 |
| PDGF | 0.000(0.153) | 1.455(0.100) | 63.039 | 1/89 | <0.001 |
| VEGF | 0.180(0.188) | -0.129(0.123) | 1.890 | 1/89 | 0.173 |
| HGF | -0.003(0.166) | 1.362(0.108) | 47.535 | 1/89 | <0.001 |

CC: C-C motif chemokine ligand, CXCL: C-X-C motif chemokine ligand, LIF: leukemia inhibitory factor, MCSF: macrophage colony-stimulating factor, GCSF: granulocyte CSF, GMCSF: granulocyte-macrophage CSF, SCF: stem cell factor, SCGF: stem cell growth factor, SDF1: stromal cell-derived factor 1, NGF: nerve growth factor, PDGFB: platelet-derived growth factor, VEGF: vascular endothelial growth factor, HGF: hepatocyte growth factor.

All results of GLM analysis with age, sex, body mass index, smoking, and previous mild COVID-19 (entered as dummy) as covariates

**ESF, Table 6**. Differences in immune profiles between drug-naive and antidepressant-treated, first-episode major dysmood disordered (FE-MDMD) patients.

| **Variables**  **(z scores)** | **Antidepressant-treated FE-MDMD (n=40)** | **Drug-naive FE-MDMD (n=71)** | **F/X2** | **df** | **p-value** |
| --- | --- | --- | --- | --- | --- |
| Classical M1 | 0.825(0.145) | 1.023(0.176) | 0.732 | 1/57 | 0.396 |
| Alternative M2 | 0.557(0.157) | 0.956(0.191) | 2.539 | 1/57 | 0.117 |
| z M1 – z M2 | 0.268(0.091) | 0.067(0.111) | 1.917 | 1/57 | 0.172 |
| Th-1 | 0.757(0.125) | 1.081(0.152) | 2.666 | 1/57 | 0.108 |
| Th-2 | 0.408(0.150) | 0.876(0.182) | 3.821 | 1/57 | 0.056 |
| z Th1 – z Th2 | 0.348(0.076) | 0.206(0.092) | 1.395 | 1/57 | 0.243 |
| Th-17 | 0.395(0.116) | 0.872(0.141) | 6.626 | 1/57 | 0.013 |
| IRS | 0.812(0.129) | 1.149(0.157) | 2.700 | 1/57 | 0.106 |
| CIRS | 0.586(0.141) | 1.225(0.171) | 8.124 | 1/57 | 0.006 |
| z IRS – z CIRS | 0.536(0.157) | -0.182(0.190) | 8.284 | 1/57 | 0.006 |
| Neurotoxicity | 0.691(0.128) | 1.113(0.155) | 4.302 | 1/57 | 0.043 |

M: macrophage, Th: T helper, IRS: immune-inflammatory response system, CIRS: compensatory immunoregulatory system

The data are expressed as z scores (with the mean z scores of healthy controls set at 0).

**ESF, Table 7**. Differences in cytokines between drug-naive and antidepressant-treated, first-episode major dysmood disorder (FE-MDMD) patients.

| **Variables** | **Antidepressant-treated**  **FE-MDMD (n=44)** | **Drug Naive**  **FE-MDMD (n=27)** | **F/X^2^** | **Df** | **p-value** |
| --- | --- | --- | --- | --- | --- |
| IL-1α | -0.339(0.147) | 0.217(0.178) | 5.669 | 1/57 | 0.021 |
| IL-1β | -0.306(0.168) | -0.046(0.204) | 0.949 | 1/57 | 0.334 |
| IL-1RA | 0.767(0.154) | 1.144(0.187) | 2.363 | 1/57 | 0.130 |
| IL-2 | -0.237(0.159) | 0.175(0.193) | 2.650 | 1/57 | 0.109 |
| IL-2R | 0.975(0.129) | 1.392(0.156) | 4.124 | 1/57 | 0.047 |
| IFN-γ | 0.251(0.171) | 0.296(0.208) | 0.028 | 1/57 | 0.868 |
| IL-4 | 0.184(0.136) | 0.795(0.165) | 7.992 | 1/57 | 0.006 |
| IL-5 | -0.263(0.166) | -0.378(0.201) | 0.193 | 1/57 | 0.662 |
| IL-6 | 0.496(0.133) | 0.915(0.162) | 3.884 | 1/57 | 0.054 |
| IL-8 | -0.154(0.140) | 0.700(0.170) | 14.678 | 1/57 | <0.001 |
| IL-9 | 0.870(0.125) | 1.137(0.152) | 1.808 | 1/57 | 0.184 |
| IL-10 | -0.171(0.172) | 0.338(0.209) | 3.443 | 1/57 | 0.069 |
| IL-12p70 | 0.463(0.157) | 0.227(0.191) | 0.892 | 1/57 | 0.349 |
| IL-13 | 0.373(0.160) | 0.388(0.195) | 0.004 | 1/57 | 0.952 |
| IL-15 | 0.394(0.176) | 0.301(0.213) | 0.110 | 1/57 | 0.741 |
| IL-16 | 1.555(0.093) | 1.776(0.113) | 2.231 | 1/57 | 0.141 |
| IL-18 | 0.969(0.166) | 0.971(0.202) | 0.000 | 1/57 | 0.993 |
| MIF-1 | 0.143(0.118) | 0.414(0.143) | 2.099 | 1/57 | 0.153 |
| TNF-α | 0.584(0.146) | 0.911(0.177) | 1.981 | 1/57 | 0.165 |
| TNF-β | 0.984(0.132) | 1.077(0.161) | 0.197 | 1/57 | 0.659 |
| TRAIL | 1.394(0.123) | 1.535(0.150) | 0.516 | 1/57 | 0.475 |

IL: interleukin, IFN: interferon, IL1RA: interleukin receptor antagonist, MIF1: macrophage migration inhibitory factor-like protein, TNF: tumor necrosis factor, TRAIL: TNF-related apoptosis-inducing ligand.

The data are expressed as z scores (with the mean z scores of healthy controls set at 0).

**ESF, Table 8**. Differences in chemokine and growth factor levels between drug-naive and antidepressant-treated first-episode major dysmood disorder (FE-MDMD) patients.

| **Variables** | **Antidepressant-treated**  **FE-MDMD (n=44)** | **Drug Naive**  **FE-MDMD (n=27)** | **F/X^2^** | **df** | **p-value** |
| --- | --- | --- | --- | --- | --- |
| CCL2 | 0.697(0.170) | 0.810(0.207) | 0.174 | 1/57 | 0.678 |
| CCL3 | -0.459(0.136) | 0.104(0.165) | 6.746 | 1/57 | 0.012 |
| CCL4 | 0.924(0.135) | 1.063(0.164) | 0.417 | 1/57 | 0.521 |
| CCL5 | 1.144(0.100) | 1.171(0.122) | 0.028 | 1/57 | 0.868 |
| CCL7 | 0.030(0.161) | 0.723(0.196) | 7.284 | 1/57 | 0.009 |
| CCL11 | 0.519(0.164) | 0.978(0.200) | 3.077 | 1/57 | 0.085 |
| CCL27 | 1.409(0.119) | 1.439(0.144) | 0.024 | 1/57 | 0.876 |
| CXCL1 | 0.670(0.140) | 0.813(0.170) | 0.406 | 1/57 | 0.527 |
| CXCL9 | 0.617(0.154) | 0.620(0.187) | 0.000 | 1/57 | 0.990 |
| CXCL10 | 0.681(0.162) | 1.001(0.197) | 1.531 | 1/57 | 0.221 |
| LIF | 0.216(0.135) | 0.408(0.164) | 0.799 | 1/57 | 0.375 |
| M-CSF | 1.350(0.129) | 1.477(0.157) | 0.381 | 1/57 | 0.539 |
| G-CSF | -0.257(0.135) | 0.333(0.163) | 7.588 | 1/57 | 0.008 |
| GM-CSF | 0.487(0.139) | 0.567(0.168) | 0.130 | 1/57 | 0.720 |
| SCF | 0.903(0.129) | 0.900(0.157) | 0.000 | 1/57 | 0.987 |
| SCGF | 1.606(0.129) | 1.539(0.156) | 0.108 | 1/57 | 0.744 |
| SDF1 | 0.865(0.138) | 1.291(0.167) | 3.781 | 1/57 | 0.057 |
| NGF | 0.349(0.148) | 0.783(0.180) | 3.392 | 1/57 | 0.071 |
| PDGFB | 1.402(0.110) | 1.343(0.134) | 0.111 | 1/57 | 0.740 |
| VEGF | -0.183(0.165) | -0.191(0.200) | 0.001 | 1/57 | 0.977 |
| HGF | 1.193(0.134) | 1.524(0.163) | 2.397 | 1/57 | 0.127 |

CC: C-C motif chemokine ligand, CXCL: C-X-C motif chemokine ligand, LIF: leukemia inhibitory factor, MCSF: macrophage colony-stimulating actor, GCSF: granulocyte CSF, GMCSF: granulocyte-macrophage CSF, SCF: stem cell factor, SCGF: stem cell growth factor, SDF1: stromal cell-derived factor 1, NGF: nerve growth factor, PDGFB: platelet-derived growth factor, VEGF: vascular endothelial growth factor, HGF: hepatocyte growth factor.

The data are expressed as z scores (with the mean z scores of healthy controls set at 0).
